# Universal health coverage in the context of population ageing: Catastrophic health expenditure and unmet need for healthcare

**DOI:** 10.1101/2023.02.13.23285836

**Authors:** Shohei Okamoto, Mizuki Sata, Megumi Rosenberg, Natsuko Nakagoshi, Kazuki Kamimura, Kohei Komamura, Erika Kobayashi, Junko Sano, Yuzuki Hirazawa, Tomonori Okamura, Hiroyasu Iso

## Abstract

**Objectives:** This study investigates the incidence/prevalence, determinants, and consequences of catastrophic health expenditure (CHE) and unmet need for healthcare and assesses the potential heterogeneity between younger (≤ 64 years) and older people (65 years≤).

**Methods:** Utilising an annual nationally representative survey of Japanese aged 20 years and over, we estimated the incidence of CHE and unmet need for healthcare using disaggregated estimates by household members’ age (i.e. ≤ 64 years vs. 65 years≤) between 2004–2020. Using a fixed-effects model, we assessed the determinants of CHE and unmet need along with the consequences of CHE. We also assessed the heterogeneity by age.

**Results:** Households with older members were more likely to have their healthcare needs met but experienced CHE more so than households without older members. The financial consequences of CHE were heterogeneous by age, suggesting that households with older members responded to CHE by reducing food and social expenditures more so than households without older members reducing expenditure on education. Households without older members experienced an income decline in the year following the occurrence of CHE, while this was not found among households with older members. A U-shaped relationship was observed between age and the probability of experiencing unmet healthcare need.

**Conclusions:** Households with older members are more likely to experience CHE with different financial consequences compared to those with younger members. Unmet need for healthcare is more common among younger and older members than among their middle-aged counterparts, though the health consequences of this unmet need could not be determined.

## 1. Introduction

### 1.1 Universal health coverage in the context of population ageing

The achievement of universal health coverage (UHC) under the Sustainable Development Goals (SDG) requires that all people receive essential health services without being exposed to financial hardship. Previous studies have assessed global or country-level progress toward UHC in terms of essential service coverage for infants and children, antenatal care, infectious diseases, and some indicators related to non-communicable diseases, along with service capacity, access, and financial protection from large household expenditure on health, or catastrophic health expenditure (CHE) [1-3].

High-income countries show good progress according to these indicators while low- and middle-income countries still struggle to provide sufficient coverage. However, even in those countries with good service coverage and financial protection, the progress towards UHC may decelerate or be limited with respect to the growing older population for the following reasons. First, older people may seek different types of services than those included in the current global indicators used for monitoring UHC. Currently, essential health service coverage indicators are more relevant to younger populations, including common infectious diseases; reproductive, maternal, newborn, and child health matters; and non-communicable diseases [3]. Meanwhile, older people tend to need care for multiple chronic conditions, long-term care, and community support [4-9]. Second, older people are more likely to experience financial difficulties associated with health expenditure or forego care owing to cost-related barriers as they tend to suffer from chronic conditions and require frequent and intense care than younger people [10, 11]. While the former concern about service provision for older populations has been explored, the latter issue regarding the financial protection in health for older populations [11-13] needs further investigation in terms of the determinants, consequences, and required policies.

### 1.2 Measures of financial protection and healthcare access

To achieve financial protection for UHC, two things are important. First, individuals should not suffer from undue financial hardship owing to healthcare utilisation; second, regardless of their demographic and socioeconomic status, individuals should not experience an unmet need for healthcare owing to financial barriers. CHE is a concept that helps measure the level of financial hardship caused by healthcare utilisation. CHE occurs when a household’s health expenditure exceeds a certain level of capacity, assuming that the level of health expenditure goes beyond one’s ability to pay, which may reduce the expenditure on other basic needs, such as education and food. A globally standardised measure of CHE, which is also used for measuring SDG Indicator 3.8.2, defines CHE as a household’s total health expenditure beyond the 10% and 25% thresholds of the total expenditure or income [1, 14-16]. To complement the CHE measure, some studies have used impoverishing health expenditure (IHE) as an indicator to estimate the difference in the poverty headcount with and without health expenditure to capture the relative impact of CHE on household finances [17].

Healthcare access is measured through healthcare utilisation or unmet need. Under the utilisation-based approach, it is measured as an individual’s actual use of healthcare services [18-21]. Conversely, the measure of unmet need is an indicator of an individual’s lack of access to required health services. Many studies have measured unmet need based on self-assessed need for care and self-reported instances of foregone care in those situations [22-27]. Both methods of assessing healthcare access can provide biased estimations owing to heterogeneous preferences over health and healthcare across demographic and socioeconomic groups in a healthcare system with non-full access [22, 28]. However, utilisation-based measures could even be biased in the assessment of equity in healthcare access, failing to consider differences in care-seeking preferences, even when no inequity exists [22]. Although a measure for self-reported unmet need can have its limitations, self-assessed unmet need is related to poor individual health [22, 29] and population-level UHC service coverage [30]. This suggests that self-reported unmet need can be useful for measuring healthcare access and may reflect true unmet need.

### 1.3 Ageing and healthcare access: CHE and unmet need

In the human capital model (i.e. Grossman model), healthcare access is a health investment [31, 32].Individuals optimise their lifelong utility, as determined by their health status and consumption of normal goods, subject to budget and time constraints. To avoid the deterioration of health concurrent with ageing, individuals engage in healthy behaviours (e.g. healthcare utilisation and exercise). Even though healthcare utilisation is exogenous in some cases (e.g. use of emergency room owing to acute diseases such as stroke), this suggests that disparities in healthcare access arise from an individual’s observable characteristics (e.g., demographic and socioeconomic status) and unobserved heterogeneities (e.g. preference for healthcare services).

As predicted by the Grossman model, disparities in health and healthcare access across demographic and socioeconomic status can lead to heterogeneous probabilities of experiencing CHE [33-41] and unmet need [24, 42-47]. Assuming that older people need more care than younger people owing to chronic conditions, health gains (or losses) from investing (or not investing) in their health can be larger among older people. Accordingly, older people may use more care than their younger counterparts, leading to CHE. Older people or households with older family members are more likely to experience CHE than younger people or households without older members [33-36, 39-41]. Conversely, the relationship between unmet need and age is complex. By using more care, older people may meet their healthcare needs. Alternatively, they may experience unmet need because they (1) forego care owing to financial difficulties from CHE; or (2) increase the chance of forgoing care from the increased number of attempts to access healthcare services, or both. Younger people may forgo care more frequently than older people owing to their mild symptoms, high opportunity costs (i.e. workers foregoing earnings because of healthcare utilisation), and higher co-payment rates than older people in some countries (e.g. Japan). However, some studies revealed that older people tend to experience more unmet need compared to younger people [26], and others showing opposite results [43, 44, 47].

While some studies have revealed an association between unmet need and subsequent health deterioration [22, 29], evidence of the consequences of CHE remains scarce. Three potential outcomes of concern are unmet need, health consequences, and financial consequences. First, the relationships between healthcare utilisation, CHE and unmet need are complex. Greater healthcare utilisation may satisfy an individual’s need for healthcare [25], while it can also cause CHE and financial hardship that may suppress their healthcare utilisation. If an individual foregoes care because of CHE, then the subsequent health deterioration owing to unmet need will be problematic. Second, CHE can be associated with both improvement and worsening of an individual’s health. This may be related to the overuse or underuse of care and the quality of healthcare services. However, empirical challenges exist in the assessment of this issue, as individuals usually use healthcare to respond to their health issues. In short, the relationship between health and healthcare utilisation is endogenous. Moreover, owing to the nature of healthcare services, which require a high level of expertise recognised as the ‘incomplete contract’ and ‘moral hazard’ induced by the supply side [48], it is hard for non-experts to judge if they really need to receive healthcare services. However, the lack of an objective measure of individuals’ health status and data, obtained from healthcare service suppliers like health insurance claims, means that this study is unable to respond to this question. Third, CHE can affect an individual’s non-health consumption, income, and wealth through (1) financial pressure from CHE, and (2) restricted choices in these activities because of health issues. Accordingly, the heterogeneity between younger and older people is of concern. Based on the life-cycle model [49], younger people rely on their incomes, while older people, particularly after retirement, obtain their incomes mainly from pension benefits and depend on their savings. Therefore, older people are less likely to experience financial difficulties because their income is not linked to their health status. Moreover, even with CHE, older people may not suffer from financial hardship as they tend to have more savings, smaller debts, and fewer expense categories (e.g. lower expenditures for education and child support) compared to younger people [50]; therefore, they have greater ability to pay. Accordingly, the consequences of CHE and the heterogeneity across age groups must be assessed to enhance the progress monitoring of UHC in the context of population ageing.

### 1.4 Contributions of this study

This study’s focus on the heterogeneity between younger and older people expands the extant literature in three main ways. First, we assess the heterogeneity in the financial consequences of CHE between households with and without older members. Owing to differences in income sources and ability to pay between younger and older people, the financial consequences of CHE can vary for these groups. However, this has not been empirically assessed so far. If older people do not experience financial difficulties, even with CHE, then this suggests that in the context of population ageing, the progress monitoring of financial protection for UHC using the current approach may not be appropriate, and that a different, more complementary indicator may be needed. Second, we analyse the relationship between age and the probability of experiencing unmet need in greater detail. In their assessment of the heterogeneity across age groups in terms of unmet healthcare need, the previous studies have not fully considered a potential non-linear relationship between age and unmet need; instead, they have assumed a linear relationship between age and unmet need or have used an arbitrary dummy variable for age (e.g. by 10 or 20 years old) [24, 26, 43, 44, 47]. Assessing such heterogeneity is important for understanding the determinants of unmet need because both younger and older people can forego care for different reasons. Third, we analyse the association between CHE and unmet need as this has lacked empirical investigation, and the relationship remains inconclusive. While it might be assumed that lower incidence of CHE indicates stronger financial protection which improves access to health care and results in lower unmet need, it is also conceivable that lower incidence of CHE is an indication of lower utilisation of health care owing to poorer access which results in higher unmet need.

## 2. Institutional settings

To evaluate the determinants and consequences of CHE and unmet need, understanding the institutional setting is important. Accordingly, this section provides a brief review of Japan’s medical care system.

### 2.1 Healthcare insurance coverage

Under public universal health insurance, all citizens in Japan can receive one of the world’s highest levels of service coverage [51, Ministry of Health, Labour and Welfare, 52]. This coverage includes a wide range of in-kind benefits and cash benefits, such as consultations, treatments, drugs, home-visit nursing, benefits related to hospitalisation (e.g. food expenses), benefits for high-cost medical expenses, dentistry, and a childbirth lump-sum allowance. Under the employee- and community-based social insurance system, insured people pay insurance premiums, which account for half of the financing source of the healthcare insurance system. To utilise medical care services, insured people bear the cost of out-of-pocket payments (OOP) at a fixed co-payment rate of 20% for pre-school children, 30% for people aged 6–69 years, 20% for people aged 70–74 years, and 10% for people aged 75 years and over (30% for people aged 70 years or over whose income level is comparable with that of employed people). These rates are identical regardless of the insured people’s residential location. Furthermore, in many regions, medical care for newborns, children, and adolescents is subsidised by the local governments (i.e. municipalities and prefectures); therefore, citizens can use these services with OOP at below 20% of the co-payment rate or with no OOP. Although the co-payment rates are seemingly high, they account for only about 12% of healthcare financing (the remaining 40% is managed by public expenditure), meaning that individuals can use medical care at low OOP cost.

To reduce patients’ financial burden, cost containment is required through the efficient management of the medical care system. Owing to the nature of medical care services that require professional knowledge, patients typically do not have knowledge of the services that they need (i.e. a principal-agent problem with imperfect contracts). Therefore, cost containment can be achieved through supply-side control [48]. Japan’s public medical care system has three main contributors to cost containment [53]. First, according to the authors, outpatient services are the main ways for service delivery with a low level of inpatient care use, even with the high per-capita number of hospital beds for historical reason (i.e. the share of outpatient care is large even in tertiary hospitals). Second, the ubiquitous payment system, with the national uniform fee schedule for reimbursement, contributes to improving efficiency (i.e. reduced administrative costs) and equity (i.e. the same benefit package throughout Japan). The prices of drugs, devices, and services, mainly for outpatient services, are biennially revised on an item-by-item basis [54]. Third, peer-reviewed medical claims and on-site audits of medical records are conducted for cost containment and quality control. Therefore, patients’ financial burden is mitigated by curbing the costs of medical care through these supply-side controls.

### 2.2 Medical care access

Japan’s medical care system does not adopt the gatekeeping or waiting-list system used by general practitioners, meaning that patients can freely choose a clinic or a hospital for their first visit. Furthermore, there are no constraints on patients’ demands for healthcare, thus they can use healthcare services when they feel they need them. To utilise specialist care at large hospitals (e.g. university hospitals) without referral from a physician, patients must bear a certain amount of a premium fee: JPY7,000 (≈ USD50) at the initial visit and JPY2,500 (≈ USD20) for the follow-up visits. However, the fee for services is identical when patients receive the same treatment from any physician at a clinic or any specialist at a hospital. With this free access system, citizens institutionally have easy access to medical care services on demand.

### 2.3 Financial protection

Even with the low cost of OOP, patients can encounter financial difficulties from excessive OOP in instances in which they have severe or chronic health conditions and thus need intensive and continual medical care. To mitigate this, two types of financial protection are available. First, citizens can receive a tax deduction when the yearly OOP of medical care usage or over-the-counter drugs for themselves and their household members exceeds a certain amount. Second, to curb excessive financial burden, the ceiling amount for monthly OOP is determined based on an individual’s income level and age. If their OOP exceeds this level, the surplus is reimbursed or provided as in-kind benefits without OOP. When a patient reaches this level more than thrice in 12 months, the ceiling amount is reduced to lessen the financial burden. Furthermore, this financial support is available when the combined yearly costs of medical care and public long-term care become excessive; for instance, when multiple household members have long-term care needs. Some insurers provide additional financial protection by setting their own ceiling amount of medical expenditure lower than the national level.

Moreover, individuals who receive low incomes can receive financial support, such as the reduction of and exemption from insurance premiums, financial support for OOP of single-parent households who receive low incomes, and additional benefits for expenses during hospitalisation (e.g. food). Some of this support varies across insurance societies in terms of eligibility and benefit levels. Furthermore, if a citizen has no assets, no family or relative to ask for financial assistance, an extremely low income (i.e. lower than the minimum cost of living), and is incapable of working owing to disease or injury, then they are eligible for public assistance. Most of these recipients are covered by medical assistance and do not have to bear the cost of insurance premiums and OOP, as they are not enrolled in any social insurance. In sum, Japan’s medical care system provides various types of policies to ensure access to care and financial protection. Other types of financial and non-financial aids from the government are also available for those needing temporal support, including self-reliance support benefits for households with financial difficulties; injury and sickness allowance (for work-unrelated health issues among insured people of occupation-based health insurance); and postponement, reduction, or exemption of tax and social insurance premiums for other types of care than medical care (e.g. residence tax and insurance premiums for pensions and long-term care). Moreover, conditional on periods of contribution payment, individuals are eligible for disability pension benefits when they are restricted from working owing to functional limitations from health issues.

## 3. Methods

### 3.1 Data

The data for this study are from the Japan Household Panel Survey (JHPS/KHPS), an annual nationally representative household survey of Japanese aged 20 years or over. The JHPS/KHPS is a unification of the Keio Household Panel Survey (KHPS) and Japan Household Panel Survey (JHPS), which were originally separate but have many questionnaires in common, including those on household structure, individual attributes, academic background, employment status, and economic conditions. Both surveys adopt stratified two-stage random sampling that uses 24 regional and city classifications, with the number of survey subjects in each classification set in accordance with their population size as the first stage of sampling. Subsequently, the subjects are selected from basic resident registers based on designated numbers and sampling intervals as the second stage. The KHPS was first conducted in 2004, with approximately 7,000 individuals from 4,000 households, and added new samples in 2007 and 2012. The JHPS was first conducted in 2009, with 4,000 individuals, and implemented a new sample in 2019. The survey information (e.g. response rates) is available in greater detail on the project website [55].

This study analysed the unbalanced panel data during 2004–2020 because not all variables used in our analyses were measured in every wave. In a study that utilises panel data, potential bias owing to sample attrition should not be ignored. To partially address non-response bias, all analyses were weighted by inverse probabilities that responded to each subsequent wave and were estimated by logit models, conditional on respondents’ age, sex, marital status, education, employment status, and residential area at the baseline. Furthermore, we restricted our analysed sample to individuals or their spouses who were household heads, since they could provide accurate information on their households compared to other household members. This led to the final sample size of at most 65,564 observations by 7,898 unique individuals.

### 3.2 Financial hardship

To define financial hardship owing to excessive health expenditure, we used two measures, CHE and IHE, obtained from the JHPS/KHPS questionnaire on household consumption in the past month. Following the definition from previous studies and the indicator for the progress monitoring of SDG 3.8.2 [1, 14-16], CHE was defined as health expenditure beyond the 10% and 25% thresholds of total household consumption.

To complement CHE, IHE was measured as the changes in poverty headcount for equalised household income with and without out-of-pocket expenditure [17]. We used the country poverty line of each year from the national government [56] and imputed it by linear interpolation when the poverty lines were not provided by the government. In addition, to further complement our analysis, we used a continuous variable of the proportion of households’ total health expenditure among total consumption instead of CHE in some analyses.

### 3.3 Unmet need

The JHPS/KHPS asks, ‘During the past year, did you receive treatment, such as an outpatient or inpatient service?’ There are six response options: (1) ‘Did nothing as healthy’, (2) ‘Did nothing despite having symptoms’, (3) ‘Went to a hospital or clinic’, (4) ‘Hospitalised’, (5) ‘Purchased a patent medicine’, and (6) ‘Other’. To define respondents’ unmet healthcare need, we created a dichotomised variable, coded as 1 for the response, ‘Did nothing despite having symptoms’ and 0 otherwise. We excluded those who did not need healthcare services because they were healthy.

### 3.4 Empirical strategies

#### (1) Financial hardship

##### Incidence

To illustrate the number of respondents who experienced financial hardship owing to health expenditure, we first calculated the incidence of CHE and IHE in each year of the survey. The incidence of people aged 64 years or younger and people aged 65 years or over (or aged 75 years or older) was also obtained to understand the potential heterogeneity across age groups, since older people are more likely to experience financial hardship because of their increasing health needs. To measure the heterogeneity across age groups, households with at least one member aged 65 years or older were compared to households with members aged 64 years or younger.

##### Determinants

We assessed the determinants of those experiencing financial hardship by estimating a linear probability model, expressed as follows:

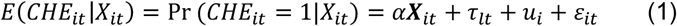

where *CHE*_*it*_ denotes the CHE status of household i in year t; ***X***_*it*_ is a vector of the independent variables, comprising a household’s demographic and socioeconomic variables; and *ε*_*it*_ is the stochastic disturbance. The independent variables ***X***_*it*_, which can affect the probability of financial hardship, comprise the following household characteristics detected in the previous studies [33-41]: Co-residing with at least one member aged 65 years or older, education (i.e. whether a household head graduated from university or higher), employment status (i.e. whether a household head was in paid employment), house ownership, the number of household members, and equalised household income (gross) and savings adjusted for the 2020 base consumer price index. Contrary to some of these studies, we did not include the household heads’ age in a model that contained a dummy variable to indicate co-residence with older people to avoid collinearity with co-residence with older family members (i.e. household heads were aged 65 years or older). When including age in the model, non-linear relationships between age and experienced CHE were considered by assessing the quadratic and cubic relationships between age and CHE. While we adopted a model with a significant coefficient for either quadratic or cubic relationships in terms of age, we used a quadratic model for the presence of neither quadratic nor cubic relationships. Income and savings, after being equalised by household size and linearly interpolated to minimise missing data, were transformed via the inverse hyperbolic sine transformation to deal with their skewness when containing a zero value [57]. We did not include health-related variables because (1) the survey did not include rich health information on household members other than respondents, and (2) by assuming that older age may be associated with worse health, we may have missed findings on age heterogeneity by controlling for household members’ health status, particularly when this is a consequence of older age.

We also controlled for city-by-year fixed-effects (*τ*_*lt*_) and individual fixed-effects (*u*_*i*_). This allowed us to control for unobserved factors that occurred in city l in year t (e.g. policy changes, economic cycle,geographical location, infrastructure, and healthcare resources) and unobserved time-invariant heterogeneity across individuals (e.g. sex of respondents and preference for healthcare use). To control for high-dimensional fixed-effects, we conducted estimations using a Stata command: *reghdfe* [58]. To address potential correlations within individuals, we also estimated cluster robust standard errors. To better understand how the nature of CHE could differ according to the age of household members, we conducted a separate analysis of households with and without a member aged 65 years or older.

##### Consequences

Even though older people may experience financial hardship owing to frequent health expenditure than their younger counterparts, the financial consequences can differ because of the variation in both ability to pay and consumption patterns between younger and older people [49]. Therefore, we investigated the heterogeneity between households with and without an older member to reveal how excessive health expenditure puts pressure on other non-health household expenditures (i.e. food, culture and recreation, social relationships, and education). Furthermore, we evaluated the financial consequences of financial hardship as follows:

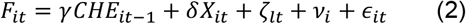

where the association of CHE for household i in year t-1 (*CHE*_*it-1*_) with financial outcomes in year t (*F*_*it*_) is assessed, controlling for independent variables and the same fixed-effects as those in Equation (1). For financial outcomes, we used income and savings (as defined earlier). We further evaluated the potential heterogeneity between households with and without an older member.

#### (2) Unmet need

##### Prevalence

We estimated the individual-level prevalence of respondents experiencing unmet need in addition to the heterogeneity between younger and older people. As unmet need was measured with individuals as the unit, respondents who were aged 65 years or older were compared to respondents who were 20–64 years.

##### Determinants

To assess the determinants of unmet need, it was important to include *need variables*, which directly affect an individual’s health need, and *non-need variables*, which indirectly affect their need [59]. Accordingly, the probability of reporting unmet need is expressed as:

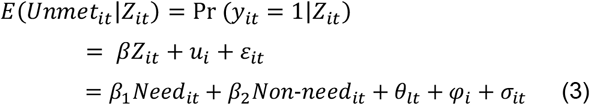

where *Need*_*it*_ denotes the need variables of individual i in year t, and *Non − need*_*it*_ represents the non-need variables. In practice, distinguishing between need and non-need variables is not easy, since individual characteristics, such as demographic factors, can be regarded as both types of variables. For example, chronological age may be viewed as a non-need variable, while older age may be regarded as a factor that influences health status. To cover a wide range of factors, we included the respondents’ characteristics as the following demographic, socioeconomic, and health-related variables identified as determinants of unmet need by the previous studies [26, 42-44, 46, 47]: Age, marital status, employment status, house ownership, household size, equalised household income and savings adjusted for the 2020-base price, self-rated health, smoking status (i.e. current smoker or not), alcohol consumption (i.e. current drinker or not), and per-week days of exercise. Non-linear relationships between age and experienced unmet need were considered in the same way as in the analysis of CHE. The model was estimated using a linear probability model, with additional controls for city-by-year fixed-effects and individual fixed-effects. Controlling for these fixed-effects was important because they could include unobserved need and non-need variables, such as national-level policy changes, access to healthcare facilities at the city level, health status of individuals before joining the survey, and individual preferences for healthcare.

The aim of this analysis was to answer two questions: (1) whether the probability of experiencing unmet need differed between younger and older people, and (2) whether the determinants of unmet need differed between younger and older people. To assess the heterogeneity across age groups, we compared the results as estimated from the respondents aged 64 years or younger with those aged 65 years or older.

#### (3) CHE and unmet need

Excessive levels of health expenditure can make individuals forgo healthcare utilisation owing to budget constraints. Conversely, their need for healthcare may be met as CHE would occur from their utilisation of healthcare. Therefore, we assessed whether CHE at wave t-1 increased or decreased the probability of experiencing unmet healthcare need at wave t using a fixed-effects linear-probability model.

## 4. Results

### 4.1 Incidence and prevalence

The incidence of CHE among the JHPS/KHPS sample was between 8.0%–12.4% at the 10% threshold and 1.1%–2.2% at the 25% threshold during the 2004–2020 period (Appendix Table A-1). Although the health expenditure data of the JHPS/KHPS were obtained as the values in the last month of the survey, which differed from the official government estimates (i.e. the monthly average in the year, calculated by the data collected every month), our estimates were comparable to the official government estimates. Furthermore, in households in which all members were aged 64 years or younger, the incidence rate was 5.2%–9.9% at the 10% threshold and 0.7%–1.7% at the 25% threshold (Appendix Table A-2). Meanwhile, the incidence was higher in households in which at least one person was aged 65 years or over, at 10.9%–22.9% at the 10% level and 1.9%–4.2% at the 25% level. Moreover, the incidence rate of IHE was around 1% for the entire sample in 2004–2018, with a higher rate in households with older people that ranged from 1.2%–4.0% (Appendix Table A-3). Conversely, the incidence rate was low among households that only had members aged 64 years or younger, at less than 1% in most years of the survey.

The prevalence rate of unmet need for healthcare moderately declined during the later periods of the survey, and ranged from 4.7%–13.1% in the entire sample (Appendix Table A-4). People aged 64 years or younger were more likely to forego healthcare compared with those aged 65 years or older; the prevalence rate was 6.2%–15.5% among younger people and 1.8%–8.6% among older people.

Table 1 shows the descriptive statistics for the household- and individual-level variables used herein. Throughout the study period, 9.8% and 7.7% of respondents had experienced CHE at the 10% threshold and unmet healthcare need, respectively. On average, the respondents spent 4.4% of their total household consumption on health expenditure. The average age of respondents or spouses (i.e. household heads) was approximately 53 years. Among the entire sample, 31.4% of the households contained at least one member aged 65 years or over. Appendix Figure A-1 presents the differences in income and savings by the presence of an older household member and CHE status. Based on the median values, households with at least one member aged 65 years or older had lower incomes but higher savings. Among households with a member aged 65 years or over, both the incomes and savings of the households experiencing CHE were slightly lower than those without CHE experience.

**Table 1.** Descriptive statistics

|  | Household-level |  |  | Individual-level |  |  |  |
| --- | --- | --- | --- | --- | --- | --- | --- |
|  | N | % or mean | SD |  | N | % or mean | SD |
| CHE10 | 65,564 | 9.8% |  | Unmet healthcare need | 34,782 | 7.6% |  |
| CHE25 | 65,564 | 1.7% |  | SRH: Very bad or bad | 34,782 | 19.8% |  |
| Total health expenditure (% of total consumption) | 65,564 | 4.4% | 0.06 | Age of respondents | 34,782 | 54.86 | 13.57 |
| Food expenditure (% of total consumption) | 65,528 | 29.4% | 0.13 | Sex of respondents: Female | 34,782 | 53.1% |  |
| Culture and recreation expenditure<br>(% of total consumption) | 65,564 | 4.3% | 0.07 | Education (respondents):<br>Bachelor+ | 34,782 | 25.0% |  |
| Social relationships expenditure<br>(% of total consumption) | 65,564 | 11.1% | 0.09 | Respondents being employed | 34,782 | 67.1% |  |
| Education expenditure (% of total consumption) | 65,564 | 1.9% | 0.04 | Marital status: Single | 34,782 | 24.1% |  |
| 65+ member living in a household | 65,564 | 31.4% |  | Current smoker | 34,782 | 19.8% |  |
| Equalised household income<br>(10,000JPY) | 65,564 | 397.89 | 281.43 | Current alcohol consumption | 34,782 | 39.5% |  |
| Equalised household savings<br>(10,000JPY) | 65,564 | 555.76 | 984.72 | Days of exercise per week | 34,782 | 0.98 | 1.76 |
| Age of household head | 65,560 | 52.86 | 14.38 |  |  |  |  |
| Education (household head): Bachelor+ | 65,564 | 33.9% |  |  |  |  |  |
| Household head being employed | 65,564 | 78.7% |  |  |  |  |  |
| House ownership | 65,564 | 80.3% |  |  |  |  |  |
| N of household members | 65,564 | 3.18 | 1.40 |  |  |  |  |

Note: SD stands for standard deviation; CHE10 and CHE25 denote catastrophic health expenditure at 10% and 25% thresholds, respectively; Expenditure for food, culture/recreation, and social relationships are equalised by household size; For household- and individual-level variables, descriptive statistics are calculated based on observations in Table 2 and Table 4, respectively.

### 4.2 Determinants

#### (1) CHE

Table 2 presents the analysis results of the determinants of CHE at the 10% and 25% thresholds. Having at least one member aged 65 years or over was associated with a higher probability of experiencing CHE at the 10% threshold (*β*: 0.02, standard error [SE]: 0.01), while the experience of CHE at the 25% threshold showed no association. Furthermore, households with higher incomes (*β*: -0.01, SE: 0.00) and household heads who had a paid occupation (*β*: -0.04, SE: 0.01) were less likely to experience CHE at the 10% threshold. The results were similar even when changing the age for a variable indicating co-residence with older household members from 65+ to 75+. CHE at the 25% threshold was associated with co-residence with a member aged 75 or older.

**Table 2.** Determinants of catastrophic health expenditure

|  | Health expenditure |  | CHE10 |  | CHE25 |  |
| --- | --- | --- | --- | --- | --- | --- |
| 65+ y/o member in a household | 0.01**<br>(0.00) |  | 0.02**<br>(0.01) |  | 0.00<br>(0.00) |  |
| 75+ y/o member in a household |  | 0.01**<br>(0.00) |  | 0.02**<br>(0.01) |  | 0.01*<br>(0.00) |
| Income | -0.00<br>(0.00) | -0.00<br>(0.00) | -0.01**<br>(0.00) | -0.01**<br>(0.00) | -0.00<br>(0.00) | 0.00<br>(0.00) |
| Savings | 0.00**<br>(0.00) | 0.00**<br>(0.00) | 0.00<br>(0.00) | 0.00<br>(0.00) | 0.00<br>(0.00) | 0.00<br>(0.00) |
| Household head being university graduate or higher | 0.00<br>(0.00) | 0.00<br>(0.00) | 0.01<br>(0.01) | 0.01<br>(0.01) | -0.00<br>(0.01) | -0.00<br>(0.01) |
| Household head being in paid work | -0.01**<br>(0.00) | -0.01**<br>(0.00) | -0.04**<br>(0.01) | -0.04**<br>(0.01) | -0.02**<br>(0.00) | -0.02**<br>(0.00) |
| House ownership | 0.01**<br>(0.00) | 0.01**<br>(0.00) | 0.03**<br>(0.01) | 0.03**<br>(0.01) | 0.01<br>(0.00) | 0.01<br>(0.00) |
| Household size | 0.00#<br>(0.00) | 0.00#<br>(0.00) | -0.00<br>(0.01) | -0.00<br>(0.01) | 0.00<br>(0.00) | 0.00<br>(0.00) |
| Individual-FE | Yes | Yes | Yes | Yes | Yes | Yes |
| City-by-Year-FE | Yes | Yes | Yes | Yes | Yes | Yes |
| Constant | 0.05**<br>(0.01) | 0.05**<br>(0.01) | 0.16**<br>(0.03) | 0.16**<br>(0.03) | 0.02*<br>(0.01) | 0.02<br>(0.01) |
| Individuals | 7,898 |  |  |  |  |  |
| Observations | 65,564 |  |  |  |  |  |

Note: Health expenditure (% of total consumption) is transformed by the inverse hyperbolic sine transformation; CHE10 and CHE25 denote catastrophic health expenditure at 10% and 25% thresholds, respectively; Estimates by fixed-effects linear probability models; \*\* p<0.01, \* p<0.05, # p<0.10; Values are coefficients with cluster-robust standard errors in parentheses; Income and savings are equalised by household size and transformed by the inverse hyperbolic sine transformation; Household size represents the log transformed number of household members; FE represents fixed-effects; Weighted by longitudinal weights to address for attrition bias; singleton observations are not used for estimations.

Table 3 shows the analysis results of the heterogeneity in the determinants of CHE by the presence of an older household member. Among the households with only members aged 64 years or younger, those with higher incomes were less likely to experience CHE. In both household types, households were less likely to experience CHE when the household heads had a paid occupation (*β*: -0.04, SE: 0.01 for both types of households). Furthermore, the association between the ages of the household heads and the probability of experiencing CHE revealed a U-shaped relationship among households without older members.

**Table 3.** Heterogeneity by the presence of an older household member: Determinants of catastrophic health expenditure

| CHE10 | With 64< | With 65+ |
| --- | --- | --- |
| Income | -0.01*<br>(0.00) | -0.01<br>(0.01) |
| Savings | 0.00<br>(0.00) | 0.00<br>(0.00) |
| Age of household head | -0.01**<br>(0.00) | -0.01<br>(0.01) |
| Age of household head<sup>2</sup> | 0.00**<br>(0.00) | 0.00<br>(0.00) |
| Household head being university graduate or higher | 0.03<br>(0.02) | -0.04<br>(0.03) |
| Household head being in paid work | -0.04**<br>(0.01) | -0.04**<br>(0.01) |
| House ownership | 0.03**<br>(0.01) | 0.08#<br>(0.05) |
| Household size | -0.01<br>(0.01) | 0.02<br>(0.02) |
| Individual-FE | Yes | Yes |
| City-by-Year-FE | Yes | Yes |
| Constant | 0.47**<br>(0.12) | 0.52<br>(0.34) |
| Individuals | 6,113 | 3,059 |
| Observations | 44,347 | 19,031 |

Note: CHE10 denotes catastrophic health expenditure at a 10% threshold; Estimates by fixed-effects linear probability models, classifying the sample by the presence/absence of at least one household member aged 65 or older; \*\* p<0.01, \* p<0.05, # p<0.10; Values are coefficients with cluster-robust standard errors in parentheses; Income and savings are equalised by household size and transformed by the inverse hyperbolic sine transformation; Household size represents the log transformed number of household members; FE represents fixed-effects; Weighted by longitudinal weights to address for attrition bias; singleton observations are not used for estimations; As a result of estimating multiple models to assess non-linear relationships between age of household head and the experienced CHE10, the quadratic relationship was observed whilst the cubic term of age was not significant.

#### (2) Unmet healthcare need

Table 4 presents the analysis results of the determinants of unmet healthcare need. We have two main findings. First, age was associated with the probability of experiencing unmet need for healthcare. Figure 1 shows this relationship and reveals a U-shaped relationship: the lowest probability of unmet need is for the age 55–60 years. Conversely, people younger and older than 55– 60 were increasingly more likely to report having forgone care. Second, only among younger people, a higher amount of savings and bad self-rated health were associated with the lower probability of experiencing unmet need while being employed was associated with the higher probability.

**Table 4.** Determinants of unmet need

|  | All | Age < 64 | 65≤Age |
| --- | --- | --- | --- |
| Age | 0.18**<br>(0.01) | 0.17**<br>(0.02) |  |
| Age<sup>2</sup> | -0.00**<br>(0.00) | -0.00<br>(0.00) | -0.00<br>(0.00) |
| Age<sup>3</sup> | 0.00**<br>(0.00) | 0.00<br>(0.00) | 0.00<br>(0.00) |
| Marital status (Single) | 0.01<br>(0.01) | 0.01<br>(0.02) | 0.01<br>(0.02) |
| Employed | 0.01#<br>(0.01) | 0.02*<br>(0.01) | 0.01<br>(0.01) |
| House ownership | -0.01<br>(0.01) | 0.00<br>(0.01) | -0.05#<br>(0.03) |
| Household size | -0.00<br>(0.01) | -0.00<br>(0.01) | -0.01<br>(0.02) |
| Income | 0.00<br>(0.00) | 0.00<br>(0.00) | 0.00<br>(0.00) |
| Savings | -0.00**<br>(0.00) | -0.00**<br>(0.00) | -0.00#<br>(0.00) |
| SRH (Bad or very bad) | -0.01*<br>(0.01) | -0.01*<br>(0.01) | -0.00<br>(0.01) |
| Current smoker | 0.03*<br>(0.01) | 0.02#<br>(0.01) | 0.06*<br>(0.02) |
| Current alcohol consumption | 0.00<br>(0.01) | 0.01<br>(0.01) | 0.01<br>(0.01) |
| Days of exercise per week | 0.00<br>(0.00) | 0.00<br>(0.00) | 0.00<br>(0.00) |
| Individual-FE | Yes | Yes | Yes |
| City-by-Year-FE | Yes | Yes | Yes |
| Constant | -8.19**<br>(0.40) | -7.07**<br>(0.38) | 3.84<br>(10.44) |
| Observations | 34,782 | 25,953 | 9,757 |
| Individuals | 5,546 | 4,419 | 1,821 |

Note: Estimates by fixed-effects linear probability models; \*\* p<0.01, \* p<0.05, # p<0.10; Values are coefficients with cluster-robust standard errors in parentheses; FE represents fixed-effects; Income and savings are equalised by household size and transformed by the inverse hyperbolic sine transformation; Household size represents the log-transformed number of household members; Weighted by longitudinal weights to address for attrition bias; singleton observations are not used for estimations.

**Figure 1.**
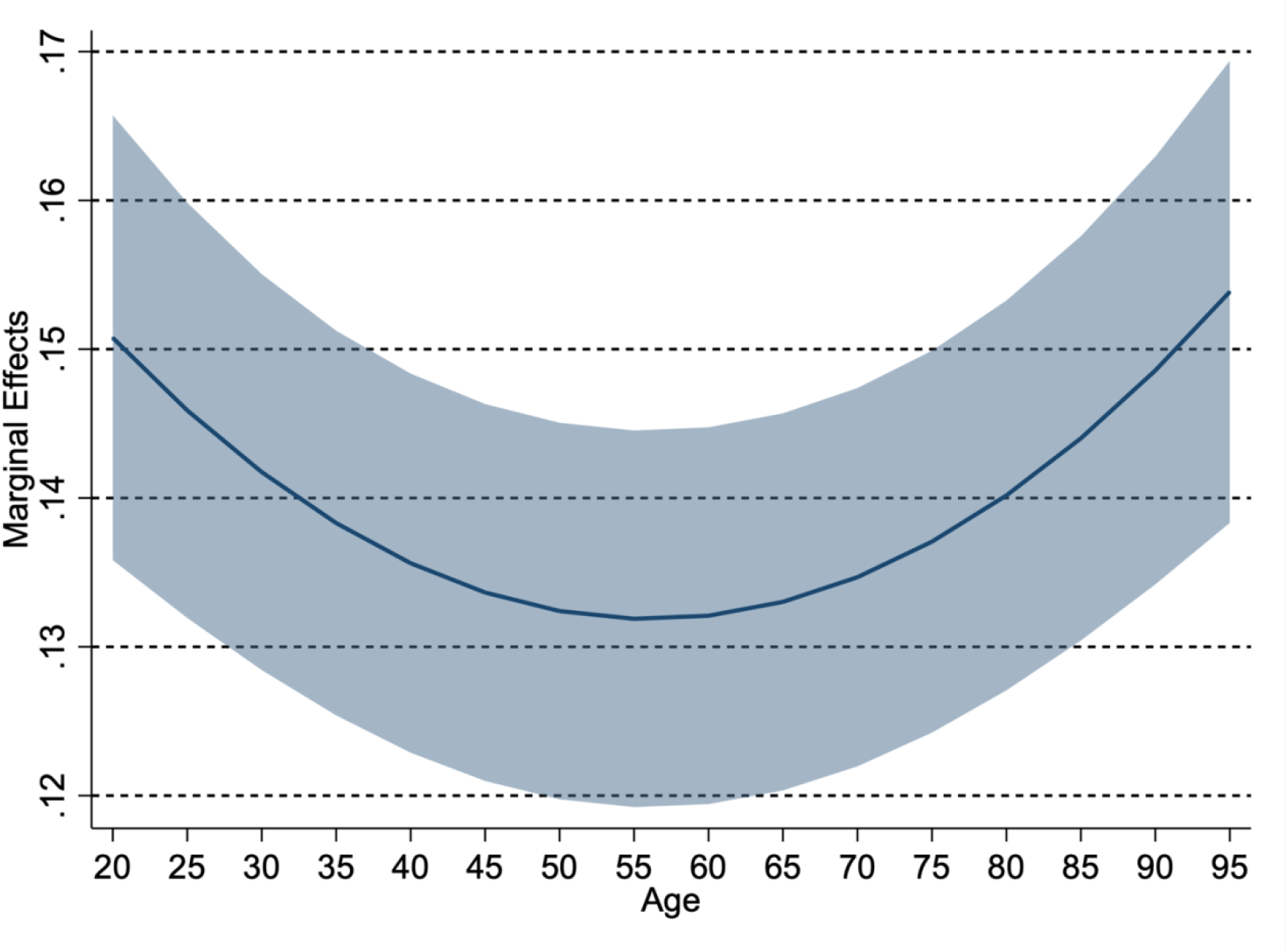
Age and unmet need for healthcare Note: The presentation of this figure is based on the estimation from the model *All* in Table 4; Line represents marginal effects of age on experienced unmet need with the shaded area representing the 95% confidence interval.

### 4.3 Consequences of CHE

#### (1) Financial consequences

Appendix Table A-5 and Figure 2 show the results of the association between CHE and food, culture or recreation, social, and education expenditures, and their heterogeneity by the presence of an older household member. CHE at 10% was associated with lower levels of food, culture or recreation, social expenditure, and education, with coefficients ranging from -0.03 to -0.01. Furthermore, reduced levels of food and social expenditure by CHE were more remarkable in households with older members (*β*: -0.01, SE: 0.00), compared to households with only members aged 64 years or younger who were more sensitive to reducing expenditure for education. Similar results were obtained even using a continuous rate of total health expenditure among total consumption.

**Figure 2.**
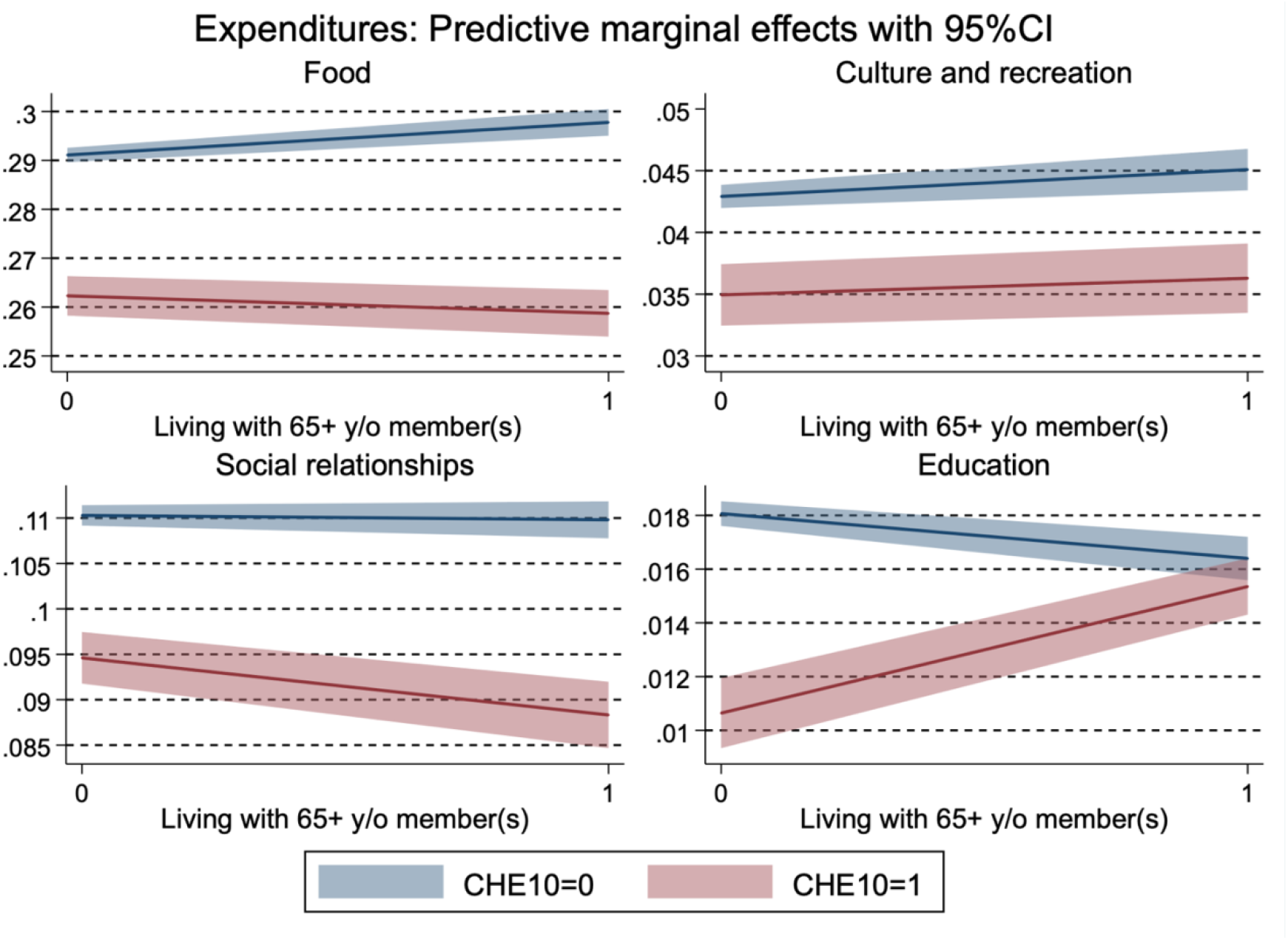
Heterogeneity by the presence of an older household member in the association between catastrophic health expenditure and food, culture/recreation, and social expenses Note: Each expenditure (% of total consumption) was transformed by the inverse hyperbolic sine transformation. Full results are presented in Appendix Table A-5.

Appendix Table A-6 and Figure 3 present the results of the financial consequences of CHE and the heterogeneity between households with and without older members. Households with only members aged 64 years or younger were more likely to experience an income decline in the following year after experiencing CHE (*β*: -0.02, SE: 0.01). Meanwhile, it was not observed among households with older members. The association between savings and CHE was not observed for both household types. With a continuous health expenditure variable, the results became less clear.

**Figure 3.**
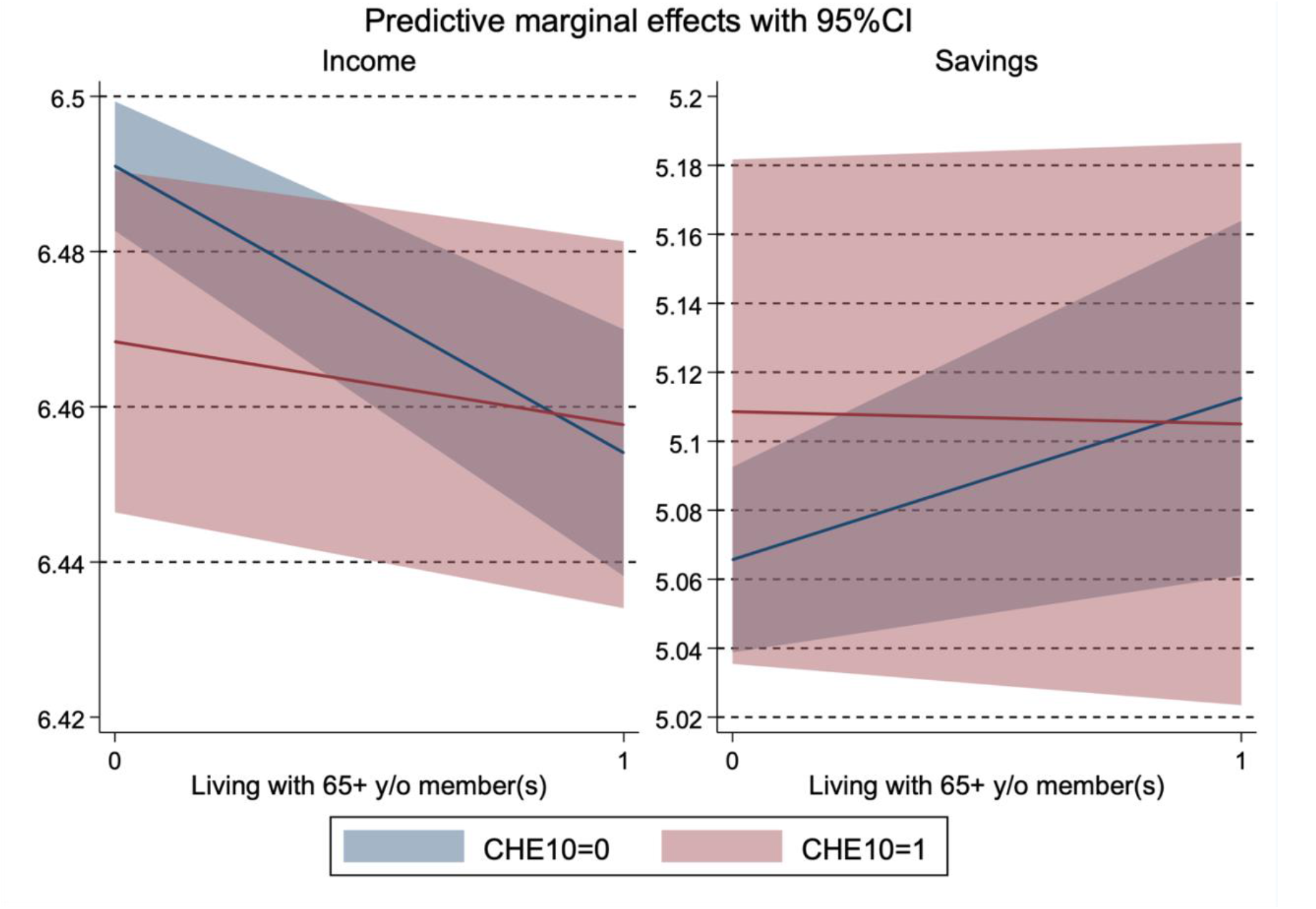
Heterogeneity by the presence of an older household member in financial consequences of catastrophic health expenditure

#### (2) CHE and unmet need

Table 5 shows the association between CHE and unmet healthcare need in the following year; no association was observed. In contrast, the continuous variable for total health expenditure was negatively associated with unmet need.

**Table 5.** Catastrophic health expenditure and unmet need

| VARIABLES | Unmet need |  |  |  |
| --- | --- | --- | --- | --- |
| CHE10[t-1] | -0.01<br>(0.00) | -0.01<br>(0.01) |  |  |
| Health expenditure [t-1] |  |  | -0.06*<br>(0.02) | -0.07#<br>(0.04) |
| Age | 0.18**<br>(0.01) | 0.13**<br>(0.01) | 0.18**<br>(0.01) | 0.13**<br>(0.01) |
| Age<sup>2</sup> | -0.00**<br>(0.00) | -0.00*<br>(0.00) | -0.00**<br>(0.00) | -0.00*<br>(0.00) |
| Age<sup>3</sup> | 0.00**<br>(0.00) | 0.00**<br>(0.00) | 0.00**<br>(0.00) | 0.00**<br>(0.00) |
| CHE10[t-1] * Age |  | 0.00<br>(0.00) |  |  |
| CHE10[t-1] * Age<sup>2</sup> |  | -0.00<br>(0.00) |  |  |
| CHE10[t-1] * Age<sup>3</sup> |  | 0.00<br>(0.00) |  |  |
| Health expenditure [t-1] * Age |  |  |  | 0.00<br>(0.00) |
| Health expenditure [t-1] * Age<sup>2</sup> |  |  |  | -0.00<br>(0.00) |
| Health expenditure [t-1] * Age<sup>3</sup> |  |  |  | 0.00<br>(0.00) |
| Observations |  | 23,458 |  |  |
| Individuals |  | 4,190 |  |  |

Note: CHE10 denotes catastrophic health expenditure at a 10% threshold; Health expenditure (% of total consumption) is mean-centralised and transformed by the inverse hyperbolic sine transformation; Estimates by fixed-effects linear probability models; \*\* p<0.01, \* p<0.05; Values are coefficients with cluster-robust standard errors in parentheses; Controlled for employment status of respondents (working or not), marital status of respondents (single or not), educational attainment of respondents (bachelor or higher), house ownership, household size, current cigarette smoking, current alcohol consumption, exercise days, individual fixed-effects, and city-by-year fixed-effects; Age of respondents is centralised when interactions with CHE were analysed; Weighted by longitudinal weights to address for attrition bias; singleton observations are not used for estimations.

## 5. Discussion

This study assesses the incidence/prevalence, determinants, and consequences of CHE and unmet healthcare need and the observable heterogeneities between younger and older people. We have five main findings. First, among households with older people, the incidence of experiencing CHE was high and unmet need was low, compared to their younger counterparts. Second, households with older members were more likely to experience CHE than households with only members aged 64 years or younger. Moreover, households with higher incomes and household heads in paid employment had a lower probability of experiencing CHE. Third, a U-shape relationship was observed between age and the likelihood of experiencing unmet need. Among younger people, being in paid employment had a higher probability of experiencing unmet need, while higher savings and negative self-rated health had a lower probability of experiencing unmet need. Fourth, after experiencing CHE, households with older people tended to reduce their expenditure on food and social relationships more so than households with only younger members. Meanwhile, households with only younger members experienced a decline in income in the following year after experiencing CHE, while households with older members did not. Fifth, there was no association between experiencing CHE and having unmet need in the following year.

Households with older members (or older people themselves) are more likely to experience CHE and have a lower prevalence of unmet need compared to households that only have younger members (or younger people). A previous study revealed that healthcare use was negatively associated with unmet need [25] because older people utilised more healthcare owing to their higher needs and their healthcare need was met. However, our analysis revealed no significant association between CHE and unmet need. As discussed in the Introduction section, CHE can affect unmet need both positively and negatively, cancelling out the overall effect. Specifically, greater healthcare use can reduce unmet need but can also result in CHE, going by our finding regarding continuous variable for total health expenditure. Conversely, greater healthcare use can cause financial difficulties, such as CHE, and lead people to forgo needed care, resulting in greater unmet need. Moreover, by needing more care, the chance of forgoing care may increase owing to the increased number of attempts to access healthcare services. In this way, it is important to try to understand the complexity and nuances that underlie the absence of a statistical relationship between CHE and unmet need. In addition, the threshold of CHE may not be appropriate to capture financial hardship.

Comparing our estimate of the incidence of CHE at 10% in Japan with estimates in other regions in 2017, it was lower than both the global and the high-income group’s medians [10]. When decomposing this into households with and without older members, both of the estimates in Japan were lower than or similar to the ones in the high-income groups. Moreover, the prevalence of unmet needs among the total and older populations was lower than the ones in most countries in the World Health Organization’s regions of Americas and the Europe [30]. Based on these indicators, Japan’s situation regarding financial protection in health may be considered better than in many other countries.

CHE resulted in different consequences between households with and without older members. Experience of CHE led both types of households to reduce their expenditure on food, culture and recreation, and social relationships. Households with older members had larger reduced expenditure on food and social relationships. There are two possible interpretations of this finding, which pose a concern about the long-term deteriorative health effects of CHE. First, owing to the large financial burden of CHE, the level of these household expenditures may be suppressed. Reduction in food expenditure leads to low energy and nutrient intake, which can increase the risk of frailty among older people [60] and eventually raise functional limitations and mortality risks [61, 62]. Second, engagement in activities related to these expenditures may be restricted because of health issues. Considering that social relationships positively affect an individual’s health status [63], this may lead to further health deterioration.

Regarding the other financial consequences of CHE (e.g. income), households with only younger members were disproportionally affected. These households saw an income decline in the year following the experience of CHE. This can be interpreted based on the Grossman model [32], assuming that the main income source of younger people is their salaries, while that of older people is their pension benefits. Accordingly, the health status of younger people is inextricably linked to their income through productivity and work hours, so their salaries may decline owing to ill health. Conversely, among older people, their health status does not affect their income when they receive pension benefits. Although we did not discover any significant results regarding savings, from the median value, households with older members had obtained more savings, which potentially suggests that older people have the better ability to pay than younger people.

We also found that the probability of experiencing unmet needs was higher among young and old people and lowest in middle-aged people; however, we were unable to explore the reason for this. The findings of previous studies are relevant here although differences in institutional settings across countries should be noted. In a Canadian survey that had a relatively young sample aged 12 years or over (average age of 48 years) wait time was the most common reason for unmet needs [22]. Meanwhile, among older people, economic hardship was the leading reason for unmet healthcare needs, with the additional finding that chronic conditions increased the probability of experiencing unmet needs [64]. These findings suggest that younger and older people may forgo care for different reasons. For younger people who are employed, the opportunity cost of seeking care can be an important reason for unmet healthcare needs. Here, households with employed heads were less likely to experience CHE, which may relate to forgone care owing to the opportunity cost. Furthermore, younger people may develop potentially milder symptoms, so they may not have actual or perceived need to use healthcare services. Conversely, older people may forgo care owing to financial hardship or accessibility issues arising from multiple chronic conditions. The group of 55 to 60-year-olds with the lowest prevalence of unmet need may be in the optimal situation where they experience less opportunity cost from seeking healthcare (as they are retired early or are in more senior/better employment situations which allow sick leave); have sufficient income, savings, or both; and are in relatively good health such that they do not have frequent need for healthcare or experience difficulties in accessing health facilities. While the heterogeneous reasons for unmet needs between younger and older people require further investigation, it is important for appropriate policies to respond to the different health needs of younger and older people. Assessing whether one’s healthcare utilisation is appropriate from the demand side is difficult owing to the nature of healthcare services. Therefore, it is preferable for patients if they can access healthcare when they feel they need it. In our analysis, negative self-rated health was associated with a lower probability of experiencing unmet healthcare needs, potentially indicating that people who perceive need are in fact able to have those needs met by the healthcare system in Japan.

### 5.1 Implications

Our findings suggest several policy implications. First, generous financial support is needed for younger households that experience CHE, particularly when they do not have high incomes and savings. By averting unmet healthcare needs through access to adequate cures or treatments, enabling people to return to their jobs is important to further avoid productivity loss because of health issues. Accordingly, various types of government aids are available in Japan. However, compared to employed people who are covered by occupation-based social insurance, those who enrol in community-based health insurance (e.g. self-employed and casual employees) are more vulnerable to income decline owing to health shocks. Therefore, the following approaches are suggested.

First, people should be encouraged to accumulate financial assets, including enrolment in private insurance, to prepare for unexpected events (that cause catastrophic expenses and income decline) through financial education and larger financial incentives (e.g. tax deduction or credit for insurance premiums). Particularly for older people, it is necessary to improve their financial capability of decumulating their financial assets, assuming that older people tend to have stable incomes and savings. Older Japanese people decumulate their wealth more slowly than predicted owing to precautionary savings and bequests [65]. With well-designed financial planning (created by a financial planner), older people may be able to better respond to high expenses.

Second, the sickness allowance coverage and other types of social insurance should be expanded to include self-employed and casual employees because these people are more likely to skip necessary care owing to unavailability of these benefits and to avoid income decline. To reduce the opportunity cost of seeking care, the utilisation of over-the-counter drugs and the enhanced availability of occupational physicians and (paid) sick leave may be also helpful.

Third, to address unmet need for healthcare among older people, it is necessary to reduce the physical barriers to accessing healthcare services. Some physical or mental functional limitations and health issues make visiting a clinic or hospital difficult for older people. Therefore, public or private sectors should consider providing home-visit or online medical care and transportation services to visit healthcare institutions.

Finally, there are many available financial protection policies, so individuals may find it difficult to identify and use a service that best fits their situation. Therefore, to enhance service usability, it is imperative to encourage the diffusion and uptake of available services (e.g. via leaflets), reduce the administrative burden of applications, and provide technical support to identify and utilise them (e.g. via consultations).

### 5.2 Limitations

This study has several limitations. First, the JHPS/KHPS contains limited information on respondents’ health status. Therefore, we were unable to assess the health consequences of CHE and unmet need. Owing to the potential heterogeneities in the consequences of CHE and reasons for unmet need between younger and older people, the health consequences may differ between these groups.

Second, the measure for unmet need should be more precise. The JHPS/KHPS asks how respondents deal with their symptoms. As such, even foregoing care owing to mild symptoms, which may not cause the subjective need for care, can be regarded as unmet need. Therefore, a measure for unmet need should reflect an individual’s need for care. Moreover, a binary measure of self-assessed unmet need can detect more unmet need among people who frequently attempt to access care [22]. Ideally, the probability of experiencing unmet need (i.e. the number of failures to access healthcare out of the number of attempts to access healthcare) should be used. However, doing so may be unrealistic owing to recall bias, so asking about the frequency of experiencing unmet need may be helpful, even though measurement error is still a concern.

Third, the CHE measure should be improved. The JHPS/KHPS contains information about monthly expenses. Even though our estimates for CHE incidence were comparable with the official estimates by utilising yearly data, yearly expenses would have been more appropriate because some of the independent variables (e.g. income and unmet healthcare need) were obtained on a yearly basis. Moreover, obtaining information on members who actually have healthcare costs may help to precisely assess the heterogeneity in CHE between younger and older people. Furthermore, a previous study has suggested that the definition of CHE used in the SDG Indicators overestimates its incidence among rich households and underestimates it among poor households [66]. Considering the potential differences in ability to pay between younger and older people, an alternative way to define CHE among older people or populations with large proportions of older people may be needed to reflect that they tend to have a larger amount of savings as well as more stable incomes from pension benefits. Also, a threshold of CHE (i.e. 10% of total consumption or income) may not be one-size-fits-all for populations with different demographic and socioeconomic characteristics and all countries with different healthcare systems and socioeconomic developments. Each country may need to reflect on their contexts to define CHE for a more efficient indicator of financial hardship, which can induce significant behavioural and financial consequences. In this regard, it would be necessary to complement the indicator to identify the consequences of financial hardship associated with healthcare utilisation.

Fourth, we utilised a linear probability model to estimate the probability of experiencing CHE and unmet need for easy interpretation and controlling for high-dimensional city-by-year fixed-effects. However, it should be noted that our estimates may be biased because of model misspecification.

Fifth, as the focus of the survey is largely on the working-age population, the JHPS/KHPS does not contain many older respondents. Moreover, respondents to the survey may be biased towards healthier or richer people. In addition, if older respondents live in a long-term care facility, they are unlikely to be included in the survey. Therefore, selection bias may be a concern, especially among older people.

Finally, given that Japan’s universal health insurance system has various available financial protection policies, our findings may not be applicable in other countries that have different socioeconomic and demographic factors, healthcare systems, and financial protection policies. In sum, the future studies can be enhanced by collecting data on respondents’ health status, using appropriate measures for CHE and unmet need, including a sample of many older people, and conducting cross-national analysis.

### 5.3 Conclusion

This study explores the incidence/prevalence, determinants, and consequences of CHE and unmet healthcare need and investigates the heterogeneity between younger and older people. By analysing the data from a household survey in Japan, we reveal that households with older members, 65 and over, are more likely to experience CHE with different financial consequences compared to households with only younger members under the age of 65. Unmet need for healthcare is more common among younger and older individuals than among their middle-aged counterparts, aged 55 to 60, though the health consequences of this unmet need could not be determined. Different types and levels of health and financial support may be required to meet the various age-related needs of individuals and households.

## Data Availability

The access to the data used in this study can be obtained via the website of the Panel Data Research Center at Keio University, found at

https://www.pdrc.keio.ac.jp/en/paneldata/datasets/jhpskhps/.

## Notes

**Funding** This study received funding from the World Health Organization Centre for Health Development (WHO Kobe Centre: K21003), which also provided technical support in reviewing an earlier draft of this manuscript.

### Competing Interest Statement

The authors have declared no competing interest.

### Funding Statement

This study received funding from the World Health Organization Centre for Health Development (WHO Kobe Centre: K21003), which also provided technical support in reviewing an earlier draft of this manuscript.

### Author Declarations

The access to the data was obtained via the website of the Panel Data Research Center at Keio University, found at https://www.pdrc.keio.ac.jp/en/paneldata/datasets/jhpskhps/.

