## Supplementary material for "Universal health coverage in the context of population ageing: Catastrophic health expenditure and unmet need for healthcare": SI

### Contents

**Appendix tables**


- Appendix Table A-1. Catastrophic health expenditure comparing JHPS/KHPS and official government estimates using different thresholds
- Appendix Table A-2. Catastrophic health expenditure by age of household members
- Appendix Table A-3. Impoverishing health expenditure by age of household members
- Appendix Table A-4. Unmet healthcare need by age of respondents
- Appendix Table A-5. Heterogeneity by the presence of an older household member in the association between catastrophic health expenditure and food, culture/recreation, and social expenditure
- Appendix Table A-6. Financial consequences of catastrophic health expenditure


**Appendix figures**


- Appendix Figure A-1. Disparities in incomes and savings by the presence of older members in a household and CHE status

Appendix Table A-1. Catastrophic health expenditure comparing JHPS/KHPS and official government estimates using different thresholds

| | JHPS/KHPS | | | National estimates | |
|---|---|---|---|---|---|
| year | 10% threshold | 25% threshold | N | 10% threshold | 25% threshold |
| 2004 | 8.9% | 1.4% | 3,433 | - | - |
| 2005 | 9.8% | 1.1% | 3,003 | - | - |
| 2006 | 10.4% | 1.5% | 2,758 | - | - |
| 2007 | 12.4% | 1.6% | 3,824 | - | - |
| 2008 | 11.2% | 1.4% | 3,500 | - | - |
| 2009 | 8.6% | 1.7% | 6,708 | - | - |
| 2010 | 9.7% | 1.3% | 6,067 | 9.1% | 1.6% |
| 2011 | 9.5% | 1.9% | 5,751 | 9.2% | 1.6% |
| 2012 | 10.3% | 2.0% | 6,215 | 9.3% | 1.7% |
| 2013 | 9.3% | 1.1% | 5,775 | 9.1% | 1.6% |
| 2014 | 9.5% | 1.8% | 5,251 | 9.1% | 1.6% |
| 2015 | 8.1% | 2.2% | 4,905 | 9.2% | 1.6% |
| 2016 | 9.4% | 1.6% | 4,617 | 9.4% | 1.6% |
| 2017 | 8.6% | 1.4% | 4,221 | 9.6% | 1.6% |
| 2018 | 8.6% | 1.6% | 3,861 | 9.7% | 1.7% |
| 2019 | 8.0% | 2.0% | 5,272 | 10.5% | 1.9% |
| 2020 | 9.9% | 2.1% | 3,368 | 10.9% | 1.8% |

Note: Weighted by cross-sectional and longitudinal weights; Official government estimates were obtained from https://www.mofa.go.jp/mofaj/gaiko/oda/sdgs/statistics/goal3.html

Appendix Table A-2. Catastrophic health expenditure by households with different age structures

| | All 64 or younger | | | At least one person, 65 or older | | |
|---|---|---|---|---|---|---|
| year | 10% threshold | 25% threshold | N | 10% threshold | 25% threshold | N |
| 2004 | 6.9% | 1.2% | 2,814 | 19.5% | 2.3% | 619 |
| 2005 | 8.5% | 0.8% | 2,424 | 15.5% | 2.2% | 579 |
| 2006 | 8.2% | 0.9% | 2,148 | 20.2% | 4.2% | 610 |
| 2007 | 9.9% | 1.0% | 2,986 | 22.9% | 4.1% | 838 |
| 2008 | 9.9% | 1.1% | 2,655 | 15.8% | 2.6% | 845 |
| 2009 | 6.0% | 1.1% | 4,761 | 14.0% | 3.1% | 1,947 |
| 2010 | 6.7% | 0.7% | 4,214 | 15.2% | 2.4% | 1,853 |
| 2011 | 7.3% | 1.4% | 3,922 | 13.8% | 2.7% | 1,829 |
| 2012 | 8.0% | 1.3% | 4,206 | 14.7% | 3.6% | 2,009 |
| 2013 | 6.5% | 0.7% | 3,779 | 14.7% | 1.9% | 1,996 |
| 2014 | 6.5% | 0.9% | 3,285 | 14.6% | 3.4% | 1,966 |
| 2015 | 5.4% | 1.5% | 2,968 | 12.8% | 3.3% | 1,937 |
| 2016 | 5.2% | 0.8% | 2,697 | 16.2% | 2.8% | 1,920 |
| 2017 | 6.3% | 1.0% | 2,729 | 14.8% | 2.5% | 1,492 |
| 2018 | 6.4% | 1.1% | 2,421 | 14.1% | 2.8% | 1,440 |
| 2019 | 5.5% | 1.0% | 3,383 | 10.9% | 3.1% | 1,889 |
| 2020 | 8.0% | 1.7% | 1,986 | 14.2% | 3.0% | 1,382 |

Note: Households were categorised by the age of the oldest co-residing family members, including respondents themselves; Weighted by cross-sectional and longitudinal weights.

Appendix Table A-3. Impoverishing health spending by households with different age structures

| | Total | | All 64 or younger | | At least one person, 65 or older | |
|---|---|---|---|---|---|---|
| year | Incidence | N | Incidence | N | Incidence | N |
| 2004 | 0.7% | 3,340 | 0.6% | 2,734 | 1.2% | 606 |
| 2005 | 0.9% | 2,804 | 0.5% | 2,288 | 2.5% | 516 |
| 2006 | 1.3% | 2,570 | 0.7% | 2,015 | 4.0% | 555 |
| 2007 | 1.1% | 3,557 | 0.7% | 2,777 | 2.7% | 780 |
| 2008 | 0.8% | 3,289 | 0.6% | 2,504 | 1.5% | 785 |
| 2009 | 1.8% | 6,198 | 1.3% | 4,438 | 3.0% | 1,760 |
| 2010 | 1.1% | 5,700 | 0.5% | 3,981 | 2.1% | 1,719 |
| 2011 | 1.0% | 5,377 | 0.3% | 3,692 | 2.2% | 1,685 |
| 2012 | 1.2% | 5,813 | 0.7% | 3,975 | 2.2% | 1,838 |
| 2013 | 0.8% | 5,416 | 0.6% | 3,589 | 1.3% | 1,827 |
| 2014 | 0.9% | 4,972 | 0.3% | 3,146 | 1.8% | 1,826 |
| 2015 | 1.0% | 4,663 | 0.7% | 2,862 | 1.3% | 1,801 |
| 2016 | 0.8% | 4,353 | 0.2% | 2,574 | 1.8% | 1,779 |
| 2017 | 1.1% | 3,873 | 0.6% | 2,547 | 2.5% | 1,326 |
| 2018 | 1.1% | 3,674 | 0.6% | 2,341 | 2.5% | 1,333 |
| 2019 | - | - | - | - | - | - |
| 2020 | - | - | - | - | - | - |

Note: Weighted by cross-sectional and longitudinal weights; In 2019 and 2020, the poverty line was not imputed because the poverty line after 2018 was not proved by the national government.

Appendix Table A-4. Unmet health care need by the age of respondent

| | Total | | 64 or younger | | 65 or older | |
|---|---|---|---|---|---|---|
| year | Proportion | N | Proportion | N | Proportion | N |
| 2004 | - | - | - | - | - | - |
| 2005 | 11.3% | 2,157 | 12.6% | 1,667 | 6.5% | 490 |
| 2006 | 13.1% | 1,963 | 15.5% | 1,476 | 5.6% | 487 |
| 2007 | - | - | - | - | - | - |
| 2008 | 10.0% | 2,414 | 10.9% | 1,728 | 7.5% | 686 |
| 2009 | 9.2% | 2,333 | 10.0% | 1,640 | 7.0% | 693 |
| 2010 | 10.0% | 2,224 | 11.4% | 1,508 | 7.0% | 716 |
| 2011 | 10.2% | 2,101 | 12.2% | 1,385 | 6.5% | 716 |
| 2012 | 9.0% | 2,698 | 9.3% | 1,782 | 8.6% | 916 |
| 2013 | 7.8% | 2,494 | 9.3% | 1,575 | 5.3% | 919 |
| 2014 | 8.7% | 4,075 | 11.7% | 2,345 | 4.7% | 1,730 |
| 2015 | 9.6% | 3,861 | 12.8% | 2,121 | 5.5% | 1,740 |
| 2016 | 9.3% | 3,631 | 11.3% | 1,917 | 7.0% | 1,714 |
| 2017 | 7.2% | 3,397 | 9.0% | 1,992 | 3.6% | 1,405 |
| 2018 | 5.2% | 3,431 | 6.5% | 2,000 | 2.6% | 1,431 |
| 2019 | 5.1% | 4,781 | 7.1% | 2,884 | 3.4% | 1,897 |
| 2020 | 4.7% | 2,997 | 6.2% | 1,650 | 1.8% | 1,347 |

Note: *Unmet needs* excludes those who did not experience forgone care because they were healthy; In 2004 and 2007, the question on unmet health need was not asked; Weighted by cross-sectional and longitudinal weights; Age group categorisation is based on age of survey respondents.

Appendix Table A-5. Heterogeneity by the presence of an older household member in the association between catastrophic health expenditure and food, culture/recreation, and social expenditure

| | Expenditures | | | | | | | | | | | | | | | |
|---|---|---|---|---|---|---|---|---|---|---|---|---|---|---|---|---|
| | Food | | | | Culture and recreation | | | | Social | | | | Education | | | |
| 65+ y/o member in a household | 0.01** | 0.01** | 0.01** | 0.01** | 0.00 | 0.00# | 0.00# | 0.00# | -0.00 | -0.00 | -0.00 | -0.00 | -0.00 | -0.00** | -0.00 | -0.00# |
| | (0.00) | (0.00) | (0.00) | (0.00) | (0.00) | (0.00) | (0.00) | (0.00) | (0.00) | (0.00) | (0.00) | (0.00) | (0.00) | (0.00) | (0.00) | (0.00) |
| CHE10 | -0.03** | -0.03** | | | -0.01** | -0.01** | | | -0.02** | -0.02** | | | -0.00** | -0.01** | | |
| | (0.00) | (0.00) | | | (0.00) | (0.00) | | | (0.00) | (0.00) | | | (0.00) | (0.00) | | |
| 65+ y/o in a household * CHE10 | | -0.01** | | | | -0.00 | | | | -0.01* | | | | 0.01** | | |
| | | (0.00) | | | | (0.00) | | | | (0.00) | | | | (0.00) | | |
| Health expenditure | | | -0.23** | -0.20** | | | -0.07** | -0.07** | | | -0.12** | -0.10** | | | -0.03** | -0.05** |
| | | | (0.01) | (0.01) | | | (0.01) | (0.01) | | | (0.01) | (0.01) | | | (0.00) | (0.00) |
| 65+ y/o in a household * Health expenditure | | | | -0.08** | | | | -0.00 | | | | -0.05** | | | | 0.05** |
| | | | | (0.02) | | | | (0.01) | | | | (0.01) | | | | (0.00) |
| Individuals | 7,892 | | | | 7,898 | | | | 7,898 | | | | 7,898 | | | |
| Observations | 65,521 | | | | 65,564 | | | | 65,564 | | | | 65,564 | | | |

Note: CHE10 denotes catastrophic health expenditure at a 10% threshold; Each expenditure (% of total consumption) is transformed by the inverse hyperbolic sine transformation; Health expenditure (% of total consumption) is mean-centralised and transformed by the inverse hyperbolic sine transformation; Estimates by fixed-effects linear models; ** p<0.01, * p<0.05, $^{\#}$ p<0.10; Values are coefficients with cluster-robust standard errors in parentheses; Controlled for income, savings, education of household head, employment status of household head, house ownership, household size, individual fixed-effects, and city-by-year fixed-effects; Weighted by longitudinal weights to address for attrition bias; singleton observations are not used for estimations.

Appendix Table A-6. Financial consequences of catastrophic health expenditure

| | Income | | | | Savings | | | |
|---|---|---|---|---|---|---|---|---|
| CHE10[t-1] | -0.01 | -0.02* | | | 0.02 | 0.04 | | |
| | (0.01) | (0.01) | | | (0.03) | (0.04) | | |
| 65+ y/o member in a household | -0.03** | -0.04** | -0.03** | -0.03** | 0.04 | 0.05 | 0.04 | 0.04 |
| | (0.01) | (0.01) | (0.01) | (0.01) | (0.04) | (0.04) | (0.04) | (0.04) |
| CHE10[t-1] * 65+ y/o member in a household | | 0.03# | | | | -0.05 | | |
| | | (0.01) | | | | (0.05) | | |
| Health expenditure[t-1] | | | -0.05 | -0.12# | | | 0.08 | 0.29 |
| | | | (0.04) | (0.06) | | | (0.14) | (0.19) |
| Health expenditure[t-1] * 65+ y/o member in a household | | | | 0.15# | | | | -0.45# |
| | | | | (0.08) | | | | (0.28) |
| Household head being university graduate or higher | 0.07* | 0.07* | 0.07* | 0.07* | -0.04 | -0.04 | -0.04 | -0.04 |
| | (0.03) | (0.03) | (0.03) | (0.03) | (0.08) | (0.08) | (0.08) | (0.08) |
| Household head being in paid work | 0.21** | 0.21** | 0.21** | 0.21** | -0.02 | -0.02 | -0.02 | -0.02 |
| | (0.02) | (0.02) | (0.02) | (0.02) | (0.04) | (0.04) | (0.04) | (0.04) |
| House ownership | 0.11** | 0.11** | 0.11** | 0.11** | -0.06 | -0.06 | -0.06 | -0.06 |
| | (0.02) | (0.02) | (0.02) | (0.02) | (0.08) | (0.08) | (0.08) | (0.08) |
| Household size | -0.19** | -0.19** | -0.19** | -0.19** | -0.35** | -0.35** | -0.35** | -0.35** |
| | (0.02) | (0.02) | (0.02) | (0.02) | (0.05) | (0.05) | (0.05) | (0.05) |
| Individual-FE | Yes | Yes | Yes | Yes | Yes | Yes | Yes | Yes |
| City-by-Year-FE | Yes | Yes | Yes | Yes | Yes | Yes | Yes | Yes |
| Constant | 6.41** | 6.41** | 6.40** | 6.40** | 5.49** | 5.49** | 5.50** | 5.50** |
| | (0.03) | (0.03) | (0.03) | (0.03) | (0.09) | (0.09) | (0.09) | (0.09) |
| Individuals | 7,010 | | | | 7,033 | | | |
| Observations | 55,233 | | | | 55,409 | | | |

Note: CHE10 denotes catastrophic health expenditure at a 10% threshold; Health expenditure (% of total consumption) is mean-centralised and transformed by the inverse hyperbolic sine transformation; Estimates by fixed-effects linear models; ** p<0.01, * p<0.05; Values are coefficients with cluster-robust standard errors in parentheses; FE represents fixed-effects; Income and savings are equalised by household size and transformed by the inverse hyperbolic sine transformation; Household size represents the log-transformed number of household members; Weighted by longitudinal weights to address for attrition bias; singleton observations are not used for estimations.

Appendix Figure A-1. Disparities in incomes and savings by households with different age structures and CHE status

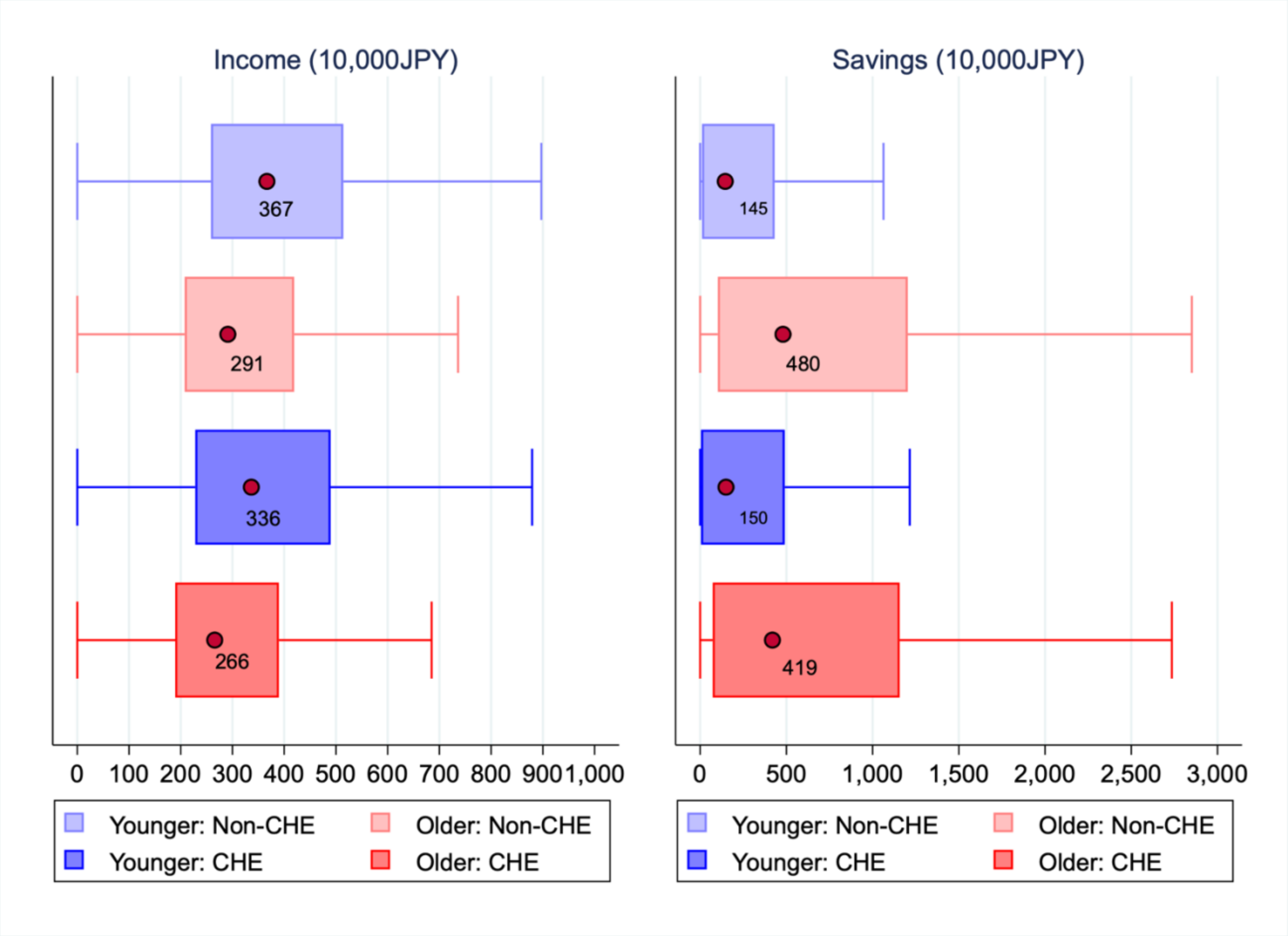


Note: 'Older' represents household with members aged 65 or older, whilst 'Younger' denotes households only with members aged < 65 years; Both income and savings are equalised by household size; Sample sizes of each category from the top are: n=42,351; n=17,409; n=3,300; n=3,152 (person-year observations, 2004-2020); Points and values in the boxes represent median values.
